# A transnational and transregional study of the impact and effectiveness of social distancing for COVID-19 mitigation

**DOI:** 10.1101/2021.09.01.21262990

**Authors:** Tarcísio M. Rocha Filho, Marcelo A. Moret, José F. F. Mendes

## Abstract

We present an analysis of the relationship between SARS-CoV-2 infection rates and a social distancing metric from data for all the states and most populous cities in the United States and Brazil, all the 22 European Economic Community countries and the United Kingdom. We discuss why the infection rate, instead of the effective reproduction number or growth rate of cases, is a proper choice to perform this analysis when considering a wide span of time. We obtain a strong Spearman’s rank order correlation between the social distancing metric and the infection rate in each locality. We show that mask mandates increase the values of Spearman’s correlation in the United States, where a mandate was adopted. We also obtain an explicit numerical relation between the infection rate and the social distancing metric defined in the present work.

## 1. Introduction

The current COVID-19 pandemic is the main health crisis in the world in a century, with over 220 million cases and 4.5 million deaths [1]. It began in China at the end of 2019, and has since expanded to every country in the world, with waves occurring at different times in each location. A number of interventions were implement in most countries, such as travel ban, social distancing and mandatory mask use [2,3], and its effects have been discussed in different works, which generally concluded that they were effective in reducing the growth of cases and deaths [4–9]. Possibly the more effective measures being lock-downs, workplaces and business closing and school closing, i. e. the social distancing policies [10], with travel restrictions expected to have modest effects in reducing transmission when there is a high circulation of the virus [11].

In order to quantify and qualify the degree of social distancing and its effects, some different approaches have been proposed: by survey questionnaires in the population in order to assess adherence to social distancing and to compare it to the growth of cases, or deaths [12], or by using mobility data from different sources [13–19]. In the latter case, a mobility or social distancing metric is compared to the growth rate of cases (or deaths) of COVID-19, or to the effective reproduction number *R*_*t*_. As we discuss below, this introduces a limitation in the analysis due to the fact that the interpretation of both the growth rate and *R*_*t*_ at the beginning of the pandemic, when most of the population is still susceptible to the virus, is different to that at latter stages, when a non-negligible proportion of the population has already been infected, of has already been vaccinated. A more informative parameter, that better represents information on the circulation of the SARS-CoV-2 virus, is the average infection rate 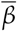, which is proportional to *R*_*t*_ divided by the proportion of the susceptible population (see Eq. 3 below). This explains particularly the result by Gatalo et al. [20] who obtained a strong Pearson correlation to *Journal Not Specified* https://www.mdpi.com/journal/notspecified between phone mobility data and COVID-19 growth rates at earlier stages, but a weaker correlation at later stages, for 25 counties in the United States.

We present here an analysis of the effect of social distancing for 22 European countries and for the 50 and 27 states of the United States and Brazil, and the most populous cities and municipalities for the latter two, respectively. These localities have different situations and histories of the pandemic. For instance, as mask use became mandatory at different moments for American states, we were able to obtain quantitative evidence on its effect on enhancing social distancing policies.

Our main goal is to evidence a monotonous relationship between social distancing data and the value of the infection rate, and to quantify it explicitly.

## 2. Material and methods

### 2.1. Effective reproduction number

The effective reproduction number *R*_*t*_(*i*) at day *i*, estimated from the generation time distribution *w*_*j*_ with *j* the number of days between infections, is given by [21]:

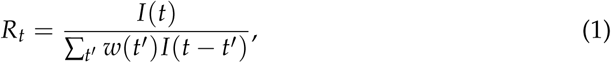

with *I*(*t*) the number (or proportion) of infected individual at day *i*. The effective reproduction number can also be estimated from the series of deaths by first determining the number of infected individuals as:

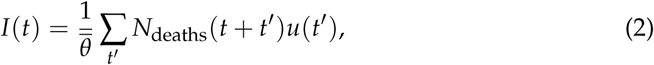

where *u*(*t*) is the distribution of the number *t* of days (taken as discrete) between first symptom and death [22], *N*_deaths_(*t*) the number of deaths at day *t* and 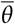 is the average infection fatality ratio [23], computed from the demographic structure in each locality. We then use Eq. (1 to determine *R*_*t*_ at a given day.

### 2.2. Infection rate

The infection rate can be estimated as [24]:

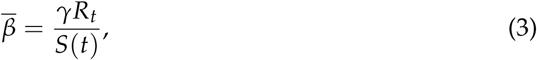

with *S*(*t*) the proportion of susceptible individuals in the population at day *t, R*_*t*_ the time dependent effective reproduction number, and *γ* the recovery rate from infection with the value reported in the literature. [25]. We can also write that

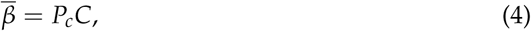

where *C* is the average number of contacts of one individual per day, and *P*_*c*_ the probability of contagion of a susceptible individual from a single contact with an infected individual. Social distancing acts by reducing the number of contacts *C*, while other non-pharmaceutical interventions reduce the value of *P*_*c*_.

### 2.3. Social distancing metric

As a proxy for the “amount” of social distancing, we define a metric quantifying the deviation from a baseline representing the pre-pandemic normality. Many possibilities exist, and different mobility data are available from different sources [26–29]. We require that data is freely available, with coverage up to the city level. For these sources only Google mobility trends satisfies these two criteria, providing data on the following six categories of locations: retail and recreation (*D*_1_); grocery and pharmacy (*D*_2_); parks (*D*_3_); transit stations (*D*_4_); workplaces (*D*_5_) and residential (*D*_6_), as percentages of variation of time spent in each type of place, with respect to a baseline defined for the period of January 3 to February 6, 2020. The symbols between parenthesis represent the numeric value of the time series for each type of data. An increase in the time spent at residence is expected to decrease the value of the infection rate 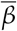, and is considered as a negative contribution to the metric, while an increase in the remaining five categories are expected to increase 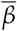 and thus contribute with a positive sign. The social distancing metric is then defined as a weighted average of the data for each category, with the specified sign, with weights given by an (arbitrarily) estimated average proportion of the duration of a day spent in each type of location, and given by

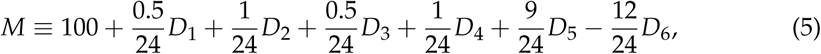

where the value of 100 is added such that the baseline is close to this value, and has no effect of the value of the Spearman’s correlation. The resulting metric *M* for each Brazilian and American state are shown in Figs. 1A and 1B, respectively, with a similar behavior for the other localities considered here (not shown). This definition is such that a smaller value of *M* represents a more beneficial situation.

**Figure 1.**
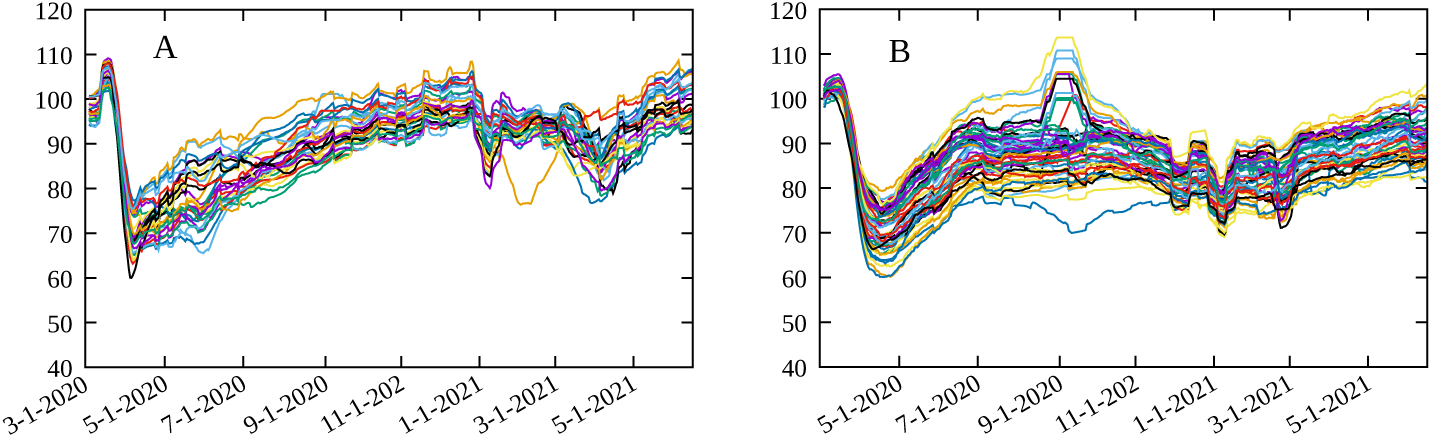
Social distancing metric *M* in Eq. (1) for A) Brazil states and B) USA states.

### 2.4. Spearman’s rank-order correlation

Spearman’s rank-order correlation *r*_*s*_(*A, B*) between two time series,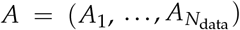 and 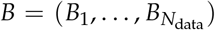, of length *N*_data_, with *A*_*i*_ the value of the series at the *i*-th data value, is defined as [30]:

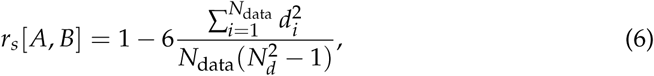

such that *−*1*≥r*_*s*_ *≥*1 and *d*_*i*_ is the difference in paired ranks of the two series *A* and *B*, i. e. the difference in position of the *i*-th data point for the two data sets when ordered in ascending order. The coefficient *r*_*s*_ measures the strength of how two variables are monotonically related, by an increasing or decreasing relation if *r*_*s*_ > 0 or *r*_*s*_ < 0, respectively.

In order to show the importance to account for the decreasing number of susceptible individuals with time, we show in Fig. 2A the time evolution of *R*_*t*_ and 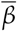 for the Los Angeles county in the United States. As the proportion of susceptible individuals decreases over time, *R*_*t*_ and 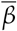 diverge slowly. By computing the Spearman’s correlation between *M* and *R*_*t*_ and between *M* and 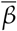, for a period of *N*_data_ = 150 days for the same data, we see from Fig. 2B that a small difference between *M* and 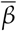 has a significant effect on the value of *r*_*s*_. The Spearman’s correlation between *M* and *R*_*t*_ is close to zero at later times while clearly positive for *M* and 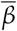. This is explained by the fact that, from Eq. (2), that the same value of *R*_*t*_ can correspond to different values of the infection rate 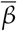 which is directly related to the circulation of the virus, as it measures the rate at which susceptible individuals are infected, and thus more closely related to the different mitigation policies implemented. We conclude that using *R*_*t*_ to represent the stage of the pandemic can lead to misleading results at later stages in assessing the effectiveness of social distancing, as the number of susceptible individuals decreases, and that of vaccinated individuals increase.

**Figure 2.**
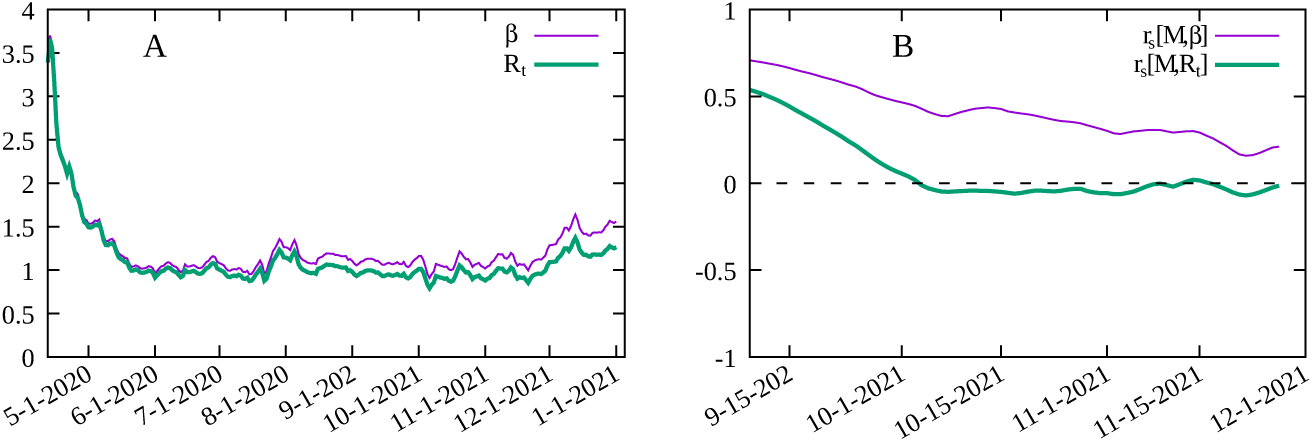
A) Time variation of 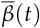 and *R*_*t*_ for Los Angeles county. B) Spearman’s rank-order correlation 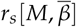 between the social distancing metric *M* and the infection rate 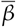 and *r*_*s*_ [*M, R*_*t*_] between *M* and the effective reproduction number *R*_*t*_ for Los Angeles county in the United States.

### 2.5. Data sources

The following data sources were employed in the present work:

- Population by age for Europe: World Population Prospects - United Nations – https://population.un.org/wpp.
- Time series of deaths and cases by country: World Health Organization – https://www.who.int/emergencies/diseases/novel-coronavirus-2019.
- Time series of cases and deaths by US counties and states: New York Times COVID- 19 Tracker data set –
- https://raw.githubusercontent.com/nytimes/covid-19-data/master/us-counties.csv.
- Population by age group in US counties and states: United States Census Bureau – https://www.census.gov/data/tables/time-series/demo/popest/2010s-counties-detail.html.
- Data on days of mask mandate in the US: Center for Disease Control and Prevention https://data.cdc.gov/Policy-Surveillance/U-S-State-and-Territorial-Public-Mask-Mandates-Fro/62d6-pm5i.
- Population by age group for Brazilian municipalities and states: Brazilian Institute for Geography and Statistics – https://brasilemsintese.ibge.gov.br/populacao.
- Time series for cases and deaths by COVID-19 by municipality and state in Brazil: Brazilian Ministry of Health – https://covid.saude.gov.br.
- Detailed data on vaccination in Brazil: Brazilian Ministry of Health – https://opendatasus.saude.gov.br/dataset/covid-19-vacinacao.

## 3. Results and discussions

The localities that are analyzed here are:

- All 50 United States states, from the first reported case up to December 20, 2021.
- The 24 United States counties with a population of at least one million and at least 1000 deaths in 2020 (Nassau was not considered due to inconsistent data for the number of deaths), from the first reported case in each county up to December 20, 2021.
- All 27 Brazilian states from February 26, 2020 to June 14, 2021.
- The 22 Brazilian cities (municipalities) with a population of at least 750 thousand from February 26, 2020 to June 14, 2021.
- All countries in the European Economic Community and the United Kingdom with Google Mobility data and at least one thousand deaths by COVID-19 in 2020, from March, 1^*st*^ 2020 to December 31, 2020, with a total of 22 countries.

The span of time of the data was chosen to avoid the effect of vaccination in the United States and Europe, while for Brazil detailed and publicly available anonimized data on each vaccine shot delivered allows modeling the time evolution of the pandemic for a longer period. For estimations of susceptible population in Eq. (3 we use the epidemiological model described in [31] to determine the attack rate in each locality and is described in Appendix A. Serological surveys also provide such estimates, but are not available for every locality and for the required time window and, where available, data do not have the required time resolution.

The results of the Spearman’s rank-order correlation between the social distancing metric *M* and the infection rate 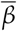 for each locality are show in Fig. 3. In order to assess the effect of mandatory mask use in each United States county and state we compute *r*_*s*_ for two periods: for the whole period, indicating in the corresponding graphic the percentage of time with a mask mandate, and for the period with a mask mandate, for those counties with a mandate for at least 50% of the days since the beginning of the pandemic, while for the remaining counties we consider the whole period and display the corresponding histogram in black. We also computed the Spearman’s correlation separately for each of the six mobility data reported by Google, with results shown in Figs. 4 and 5. The average of 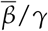, over the time period considered for each locality, versus the total number of deaths at the end of each period is shown in Fig. 6, where an approximately linear relation is clearly visible, with the exception of a few cases in Brazil.

**Figure 3.**
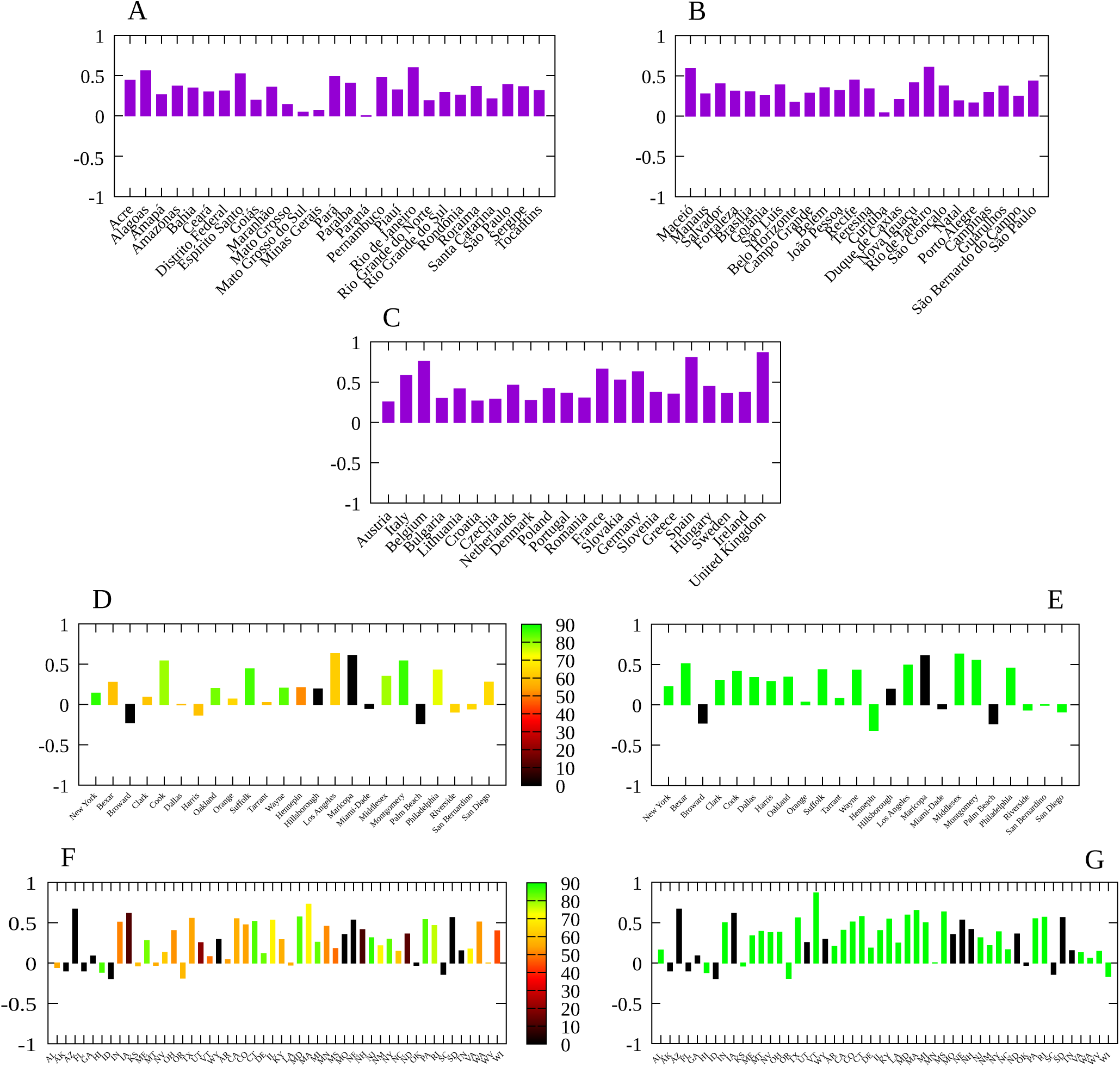
Spearman’s correlation index *r*_*s*_ between the social distancing metric *M* and the infection rate 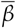 for: A) Main Brazilian municipalities with population over 750 thousand; B) Brazilian states; C) 22 European countries; d) US counties with at least one million inhabitants. D) All American states. Bar colors give the proportion of days with a mask mandate since the beginning of the pandemic in each location, up to December 20, 2020; E) Same as (D) but considering only the period with a mask mandate. States without a mask mandate in the period considered are marked in black. F) Same as (D) but for the main counties in the United states. G) Same as (E) but for the main counties in the United states.

**Figure 4.**
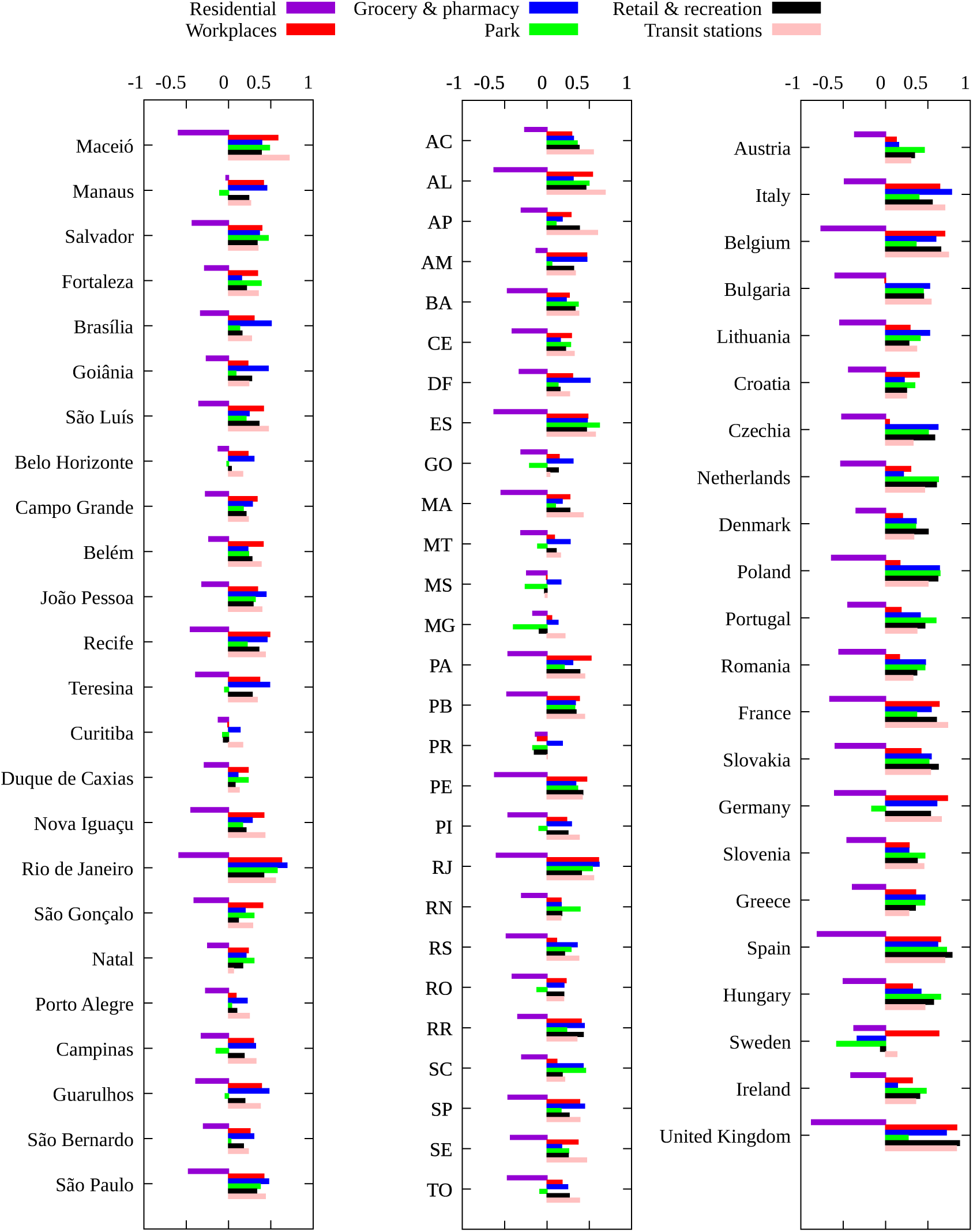
Spearman’s correlation index *r*_*s*_ between each mobility variable and the infection rate 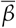 for the main Brazilian municipalities, each Brazilian state and European countries.

**Figure 5.**
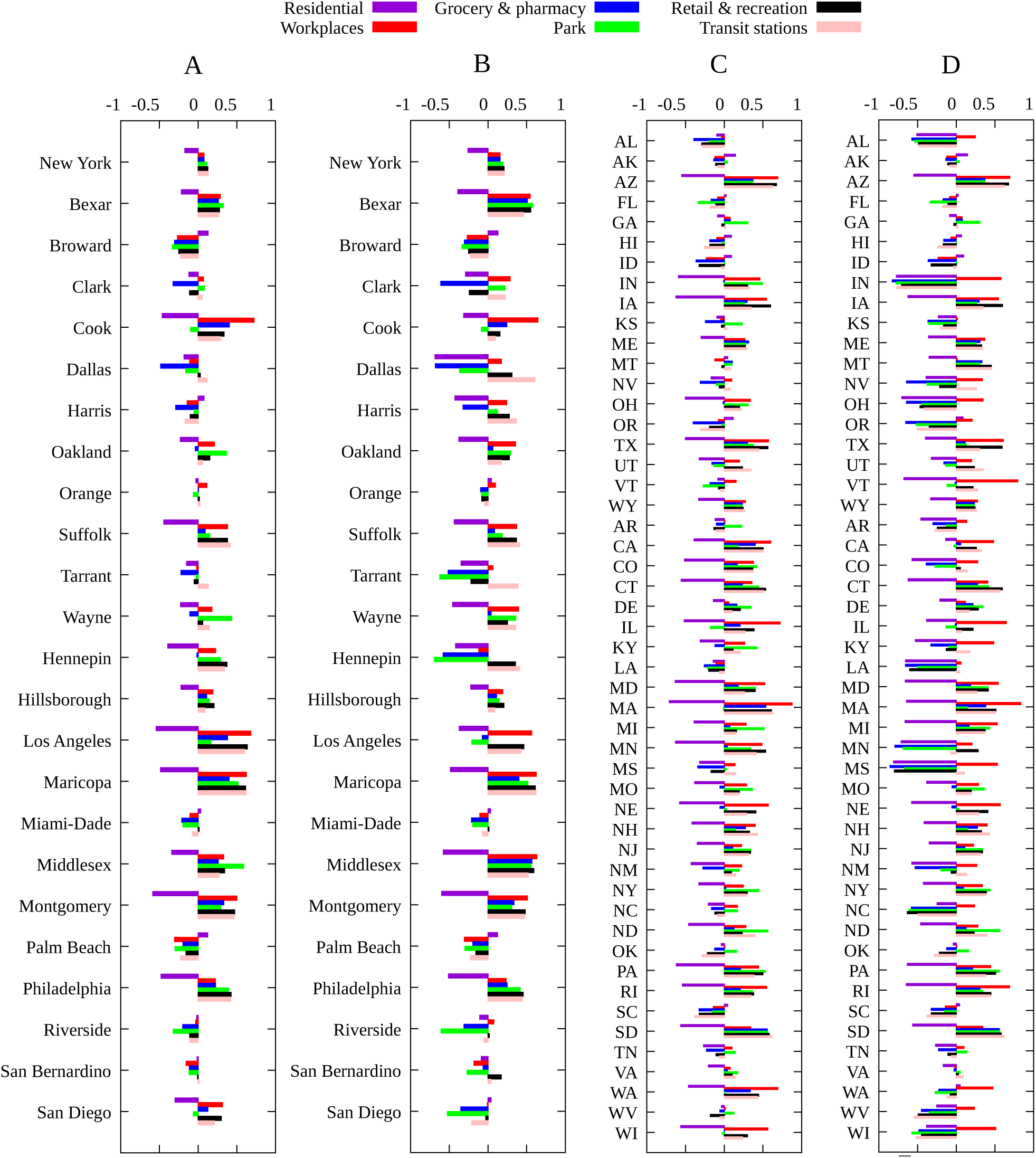
Spearman’s correlation index *r*_*s*_ between changes in each mobility category and the infection rate 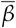 for A) Counties with more than one million inhabitants and one thousand deaths for the period from the first COVID-19 case up to December, 20 2020; B) Same as (A) but for the period with a mask mandate, except those counties with no mask mandate in 2020 (marked in black in figure 3E) for which the whole period is considered; C) Same as (A) for all US states; D) same as (B) for all US states.

**Figure 6.**
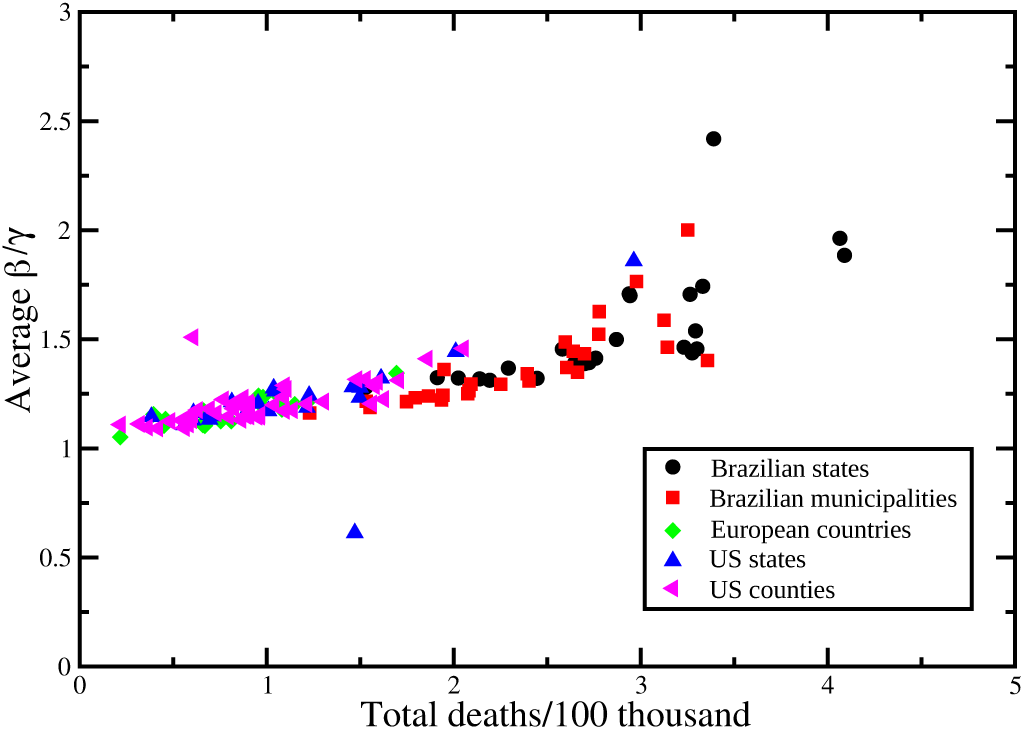
Total number of deaths per 100 thousand inhabitants at the end of the considered period as a function of the average value of 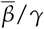during the same time span.

In order to established a numeric relationship between 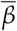 and *M* let us assume the linear relation

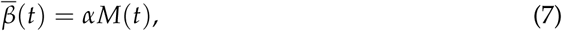

with *α* a constant, and consider only the time window that allows to an accurate estimation of *R*_*t*_. The distributions of values of the ratio 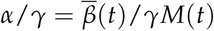 for the Brazilian states, Brazilian municipalities, European countries, United States states and counties are shown in Figs 7A–E, with values for *α*/*γ* (CI 95%) given by 0.015 (0.0096–0.023), 0.019 (0.0081–0.042), 0.014 (0.0089–0.021), 0.015 (0.0091–0.027) and 0.014 (0.0084–0.024), respectively. We also show the best fit with a log-normal distribution for values of *α*/*γ* in Figs. 7F–J.

**Figure 7.**
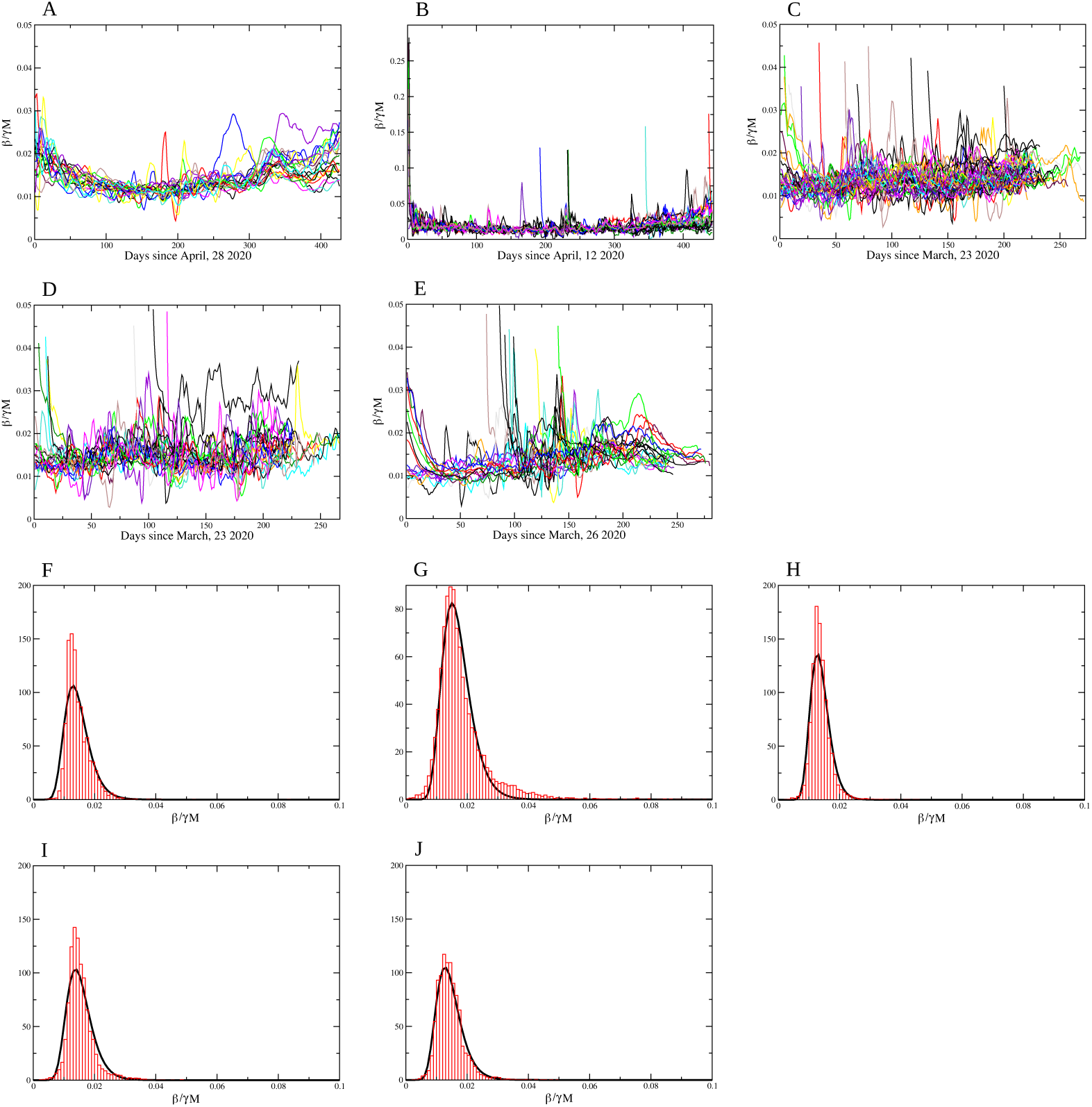
Coefficient 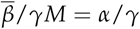 Brazilian states; B) Brazilian municipalities, C) US states; D) US counties and E) European countries. The normalized histogram (in red) and the log *−*normal distribution function (in black) for the values for *α*/*γ*: F) Brazilian states; G) Brazilian municipalities, H) US states; I) US counties and J) European countries. The values for *α*/*γ* (CI 95%) are 0.015 (0.0096–0.023), 0.019 (0.0081–0.042), 0.014 (0.0089–0.021), 0.015 (0.0091–0.027) and 0.014 (0.0084–0.024), respectively.

While vaccination reduces the proportion of susceptible individuals in the population, it does not alter the relationship of the infection rate 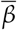 with social distancing policies with *M* as a proxy, and was explicitly taken into account in our analysis by using an epidemiological model with vaccination compartments. The approach presented here allowed to evidence a monotonous relationship between the infection rate in each locality and the social distancing metric *M*. It also allowed to explicitly obtain a numeric relationship between 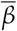 and a metric for social distancing. Behavioral changes can also have a significant impact on the evolution of any epidemic, and are difficult to include in the current analysis. Nevertheless, the significant values obtained for the Spearman’s correlation indicate the important role that social distancing has played up to now. This is particularly clear in Belgium (*r*_*s*_ = 0.75), Spain (*r*_*s*_ = 0.8) and the United Kingdom (*r*_*s*_ = 0.88), three countries with a high attack rate. The correlation is somewhat smaller for other localities, but nevertheless with significant positive values, clearly indicating an approximately monotonous relationship between the two variables.

For Brazil and the European countries the results for Spearman’s correlation are quite similar: the variation in time spent at residence is negatively correlated with the infection rate, i. e. the more time spent at home the smaller the value of 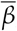, while other categories are positively correlated. For the United States, due to a much greater variety of mitigation policies implemented [13], we see a slightly different picture. In general, time at residence is negatively correlated with 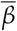 while time at workplace is positively correlated with the transmission rate, as expected. For the remaining categories (grocery and pharmacy, park, retail and recreation and transit stations) we observe both negative and positive correlations according to the locality, indicating that the most relevant categories are those related to the increase of time spent at home and the decrease of time spent at work places. For the United States case there is a significant increase in the value of *r*_*s*_ when considering only the time period with a mask mandate, which indeed shows its effectiveness.

The values of the proportionality constant *α*/*γ* between 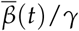 and *M*(*t*) are surprisingly close to one another, despite the great differences in the history and implemented policies to mitigate the COVID-19 pandemic. We obtain a log-normal distribution for the value of *α*/*γ* (and for *α* consequently) for all types of localities considered here, with average values significantly closer, despite all the differences between countries, implemented mitigation policies, and timings. This points to a universal efficacy of social distancing, enhanced by a mandatory mask use. The explicit linear relation in Eq. (7) with the value obtained for the proportionality constant *α* can be used, for instance, in modeling studies with different scenarios for social isolation.

Of course not only social distancing affects the evolution of the infection rate, causing the variation observed for the Spearman’s correlation for the different localities. We note that even a small increase in 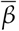, and thus a small decrease in *M*, for a long period of time, results in a significant increase in mortality, as can be seen from Fig. 6. Our analysis does not grasp the impact of great gatherings of individuals and the possible effect of the so-called superspreading events [20], or the implications of contact tracing.

## 4. Conclusion

A proper choice of a variable to represent the current circulation of the virus is central to assessing the effects of mitigation policies. The infection rate as expressed in Eq. (4) is affected by the reduction of social contacts through the average number contacts *C*, and by other implemented protocols, such as mask wearing, that reduce the probability of contagion per contact *P*_*c*_. On the other hand, the effective reproduction number *R*_*t*_, or any other measure of growth rate of the pandemic, also depends on the current attack rate, and confuses variables in the analysis. This is an important point to consider as a more detailed analysis requires a large data set, and therefore a larger time series, and therefore a significant variation in the proportion of susceptible individuals. Computing Spearman’s correlation, rather than Pearson correlation, for instance, allows us to more clearly evidence a monotonous relationship between the social distancing metric as defined here and the infection rate, and computed from the whole time series for each locality. Future research considering socioeconomic and demographic data would certainly provide valuable information on mitigation strategies targeted at specific groups, such as elders and individuals with comorbidities, as well as the impact of school closure, each considered separately from other factors [32].

We hope that the present work will contribute to a better assessment of the effects of social distancing, and at least partially of mask mandates, on the still ongoing mitigation interventions against the COVID-19 pandemic.

## Data Availability

All data used in the manuscript is publicly available and properly referred in the text.

## 5. Acknowledgments

This work received financial support from the National Council of Technological and Scientific Development - CNPq (grant number 305291/2018-1 MAM) and i3N (grant numbers UIDB/50025/2020 & UIDP/50025/2020 JFFM) - Fundação para a Ciência e Tecnologia/MEC (Portugal).

## Appendix A Epidemiological model

In order to determine the proportion of susceptible individuals in a given locality we use the approach described in [31] based on the SEIAHRV epidemiological model with variables described in Table A1

**Table A1.** Variables in the SEIAHRV model in Ref. [31]. All variables are proportions with respect to the initial population and the index *i* refers to the age-group.

| Variable | Description |
| --- | --- |
| $S_i$ | Susceptible individuals |
| $E_i$ | Exposed individuals (non-contagious) |
| $I_i$ | Infected symptomatic individuals (contagious) |
| $A_i$ | Asymptomatic symptomatic individuals (contagious) |
| $H_i$ | Hospitalized individuals |
| $V_i^{(l,k)}$ | Vaccinated individual with $l$ doses with vaccine<br>of type $k$ without primary vaccination failure |
| $U_i^{(l,k)}$ | Vaccinated individual with $l$ doses with vaccine<br>of type $k$ with primary vaccination failure |
| $R_i$ | Recovered individuals. |
| $d_l^{(i,k)}$ | individual vaccinated per unit of time with the $l$-th<br>dose of vaccine of type $k$. |
| $e_l^{(k)}$ | Efficacy of vaccine of type $k$ with $l$ doses. |

The proportion *S*_*i*_ of susceptible individuals is obtained from the epidemiological model described in [31], with model equations:

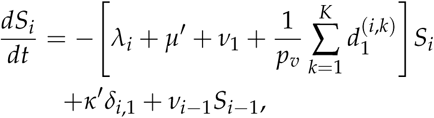

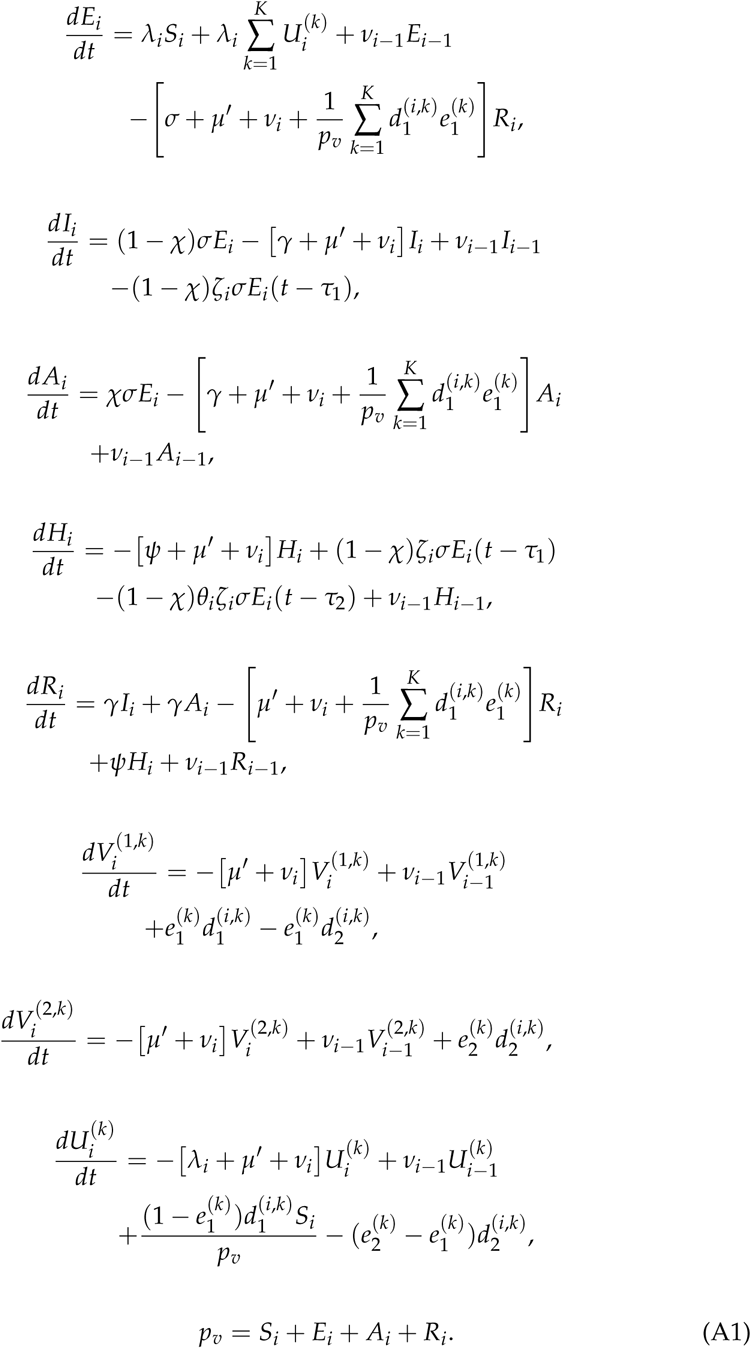

This is a non-linear delayed set of ODEs due ti the time delay between infection, hospitalization and death. The different parameter values used in the model are given in Table 1 of Ref. [31]. The force of infection in Eq. (A1) is given by

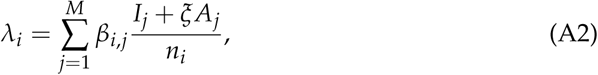

with *β*_*i,j*_) the infection rate from an infected individual of age group *j* to infect an individual of age-group *i*. The epidemiological model is calibrated using the time series of deaths in order to avoid the significant under-notification of cases [33]. The value 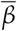 in Eq. (3) is an age-independent estimate obtained from the total proportion of susceptible individuals obtained from

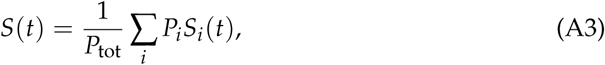

where *P*_*i*_ is the population in age group *i* and *P*_tot_ the total population for the given locality. The model is fitted from the time series of deaths as described in [31].

## References

1. John Hopkins University. John Hopkins Coronavirus Resource Center, 2021. Last accessed 4 September 2021.

2. Perra, N. Non-pharmaceutical interventions during the COVID-19 pandemic: A review. Physics Reports 2021, 913, 1. doi:10.1016/j.physrep.2021.02.001.

3. Chiesa, V.; Antony, G.; Wismar, M.; Rechel, B. COVID-19 pandemic: health impact of staying at home, social distancing and ‘lockdown’ measures - a systematic review of systematic reviews. Journal of public health (Oxf.) 2021, fdab102. Advance online publication. doi: 10.1093/pubmed/fdab102.

4. Margraf, J.; Brailovskaia, J.; Schneider, S. Adherence to behavioral COVID-19 mitigation measures strongly predicts mortality. PLoS ONE 2021, 16, e0249392. doi:10.1371/journal.pone.0249392.

5. Zhang, J.; Litvinova, M.; Liang, Y.; Wang, Y.; Wang, W.; Zhao, S. Changes in contact patterns shape the dynamics of the COVID-19 outbreak in China. Science 2021, 368, 1481. doi: 10.1126/science.abb8001.

6. Thu, T.P.B.; Ngoc, P.N.H.; Hai, N.M.; Tuan, L. Effect of the social distancing measures on the spread of COVID-19 in 10 highly infected countries. Science of The Total Environment 2020, 742, 140430. doi:10.1016/j.scitotenv.2020.140430.

7. Ayouni, I.; Maatoug, J.; Dhouib, W.; Zammit, N.; Fredj, S.B.; Ghammam, R.; Ghannem, H. Effective public health measures to mitigate the spread of COVID-19: a systematic review. BMC Public Health 2021, 21, 1015. doi:10.1186/s12889-021-11111-1.

8. Zhang, M.; Wang, S.; Hu, T.; Fu, X.; Wang, X.; Hu, Y.; Halloran, B.; Cui, Y.; Liu, H.; Liu, Z.; Bao, S. Human mobility and COVID-19 transmission: a systematic review and future directions. medRxiv preprint 2021. doi:10.1101/2021.02.02.21250889.

9. Oh, J.; Lee, H.Y.; Khuong, Q.L.; et al. Mobility restrictions were associated with reductions in COVID-19 incidence early in the pandemic: evidence from a real-time evaluation in 34 countries. Scientific Reports 2021, 11, 13717. doi:10.1038/s41598-021-92766-z.

10. Mendez-Brito, A.; Bcheraoui, C.E.; Pozo-Martina, F. Systematic review of empirical studies comparing the effectiveness of non-pharmaceutical interventions against COVID-19. Journal of Infection 2021, 83, 281. doi:10.1016/j.jinf.2021.06.018.

11. Chinazzi, M.; Davis, J.T.; Ajelli, M.; abd Marcia Litvinova, C.G.; Merler, S. y Pionti, A.P.; abd Luca ROssi, K.M.; Vespigniani, A. The effect of travel restrictions on the spread of the 2019 novel coronavirus (COVID-19) outbreak. Science 2020, 368, 395. doi:10.1126/science.aba9757.

12. Jarvis, C.I.; Van Zandvoor, K.; Gimma, A.; Prem, K.; working group, C.C..; Klepac, P.; Rubin, G.J.; Edmunds, W.J. Quantifying the impact of physical distance measures on the transmission of COVID-19 in the UK. BMC Medicine 2020, 18, 124. doi:10.1186/s12916-020-01597-8.

13. Badr, H.S.; Du, H.; Marshall, M.; Dong, E.; Squire, M.M.; Gardner, L.M. Association between mobility patterns and COVID-19 transmission in the USA: a mathematical modelling study. The Lancet Infectious Diseases 2020, 20, 1247. doi:10.1016/S1473-3099(20)30553-3.

14. Candido, D.S.; Claro, I.M.; Jesus, J.G.; Souza, W.M.; Moreira, F.R.R.; Dellicour, S. Evolution and epidemic spread of SARS-CoV-2 in Brazil. Science 2020, 369, 1255. doi:10.1126/science.abd2161.

15. Wellenius, G.A.; Vispute, S.; Espinosa, V.; Fabrikant, A.; Tsai, T.C.; Hennessy, J. Impacts of social distancing policies on mobility and COVID-19 case growth in the US. Nature Communications 2021, 12, 3118. doi:10.1038/s41467-021-23404-5.

16. Singh, B.B.; Lowerison, M.; Lewinson, R.T.; Vallerand, I.A.; Deardon, R.; Gill, J.P.S.; Sing, B.; W, B.H. Public health interventions slowed but did not halt the spread of COVID-19 in India. Transboundary and Emerging Diseases 2021, 68, 2171. doi:10.1111/tbed.13868.

17. Nouvellet, P.; Bhatia, S.; Cori, A.; Ainslie, K.E.C.; Baguelin, M.; Bhatt, S. Reduction in mobility and COVID-19 transmission. Nature Communications 2021, 12, 1090. doi:10.1038/s41467-021-21358-2.

18. Yilmazkuday, H. Stay-at-home works to fight against COVID-19: International evidence from Google mobility data. Journal of Human Behavior in the Social Environment 2021, 31, 210. doi:10.1080/10911359.2020.1845903.

19. Nova, A.C.; Ferreira, P.; Almeira, D.; Dionísio, A.; Quintino, D. Are Mobility and COVID-19 Related? A Dynamic Analysis for Portuguese Districts. Entropy 2021, 23, 786. doi: 10.3390/e23060786.

20. Gatalo, O.; Tseng, K.; Hamilton, A.; Lin, G.; Klein, E. Associations between phone mobility data and COVID-19 cases. The Lancet Infectious Diseases 2020, 21, E111. doi:10.1016/S1473-3099(20)30725-8.

21. Fraser, C. Estimating Individual and Household Reproduction Numbers in an Emerging Epidemic. PLOS One 2007, 2, e758. doi:10.1371/journal.pone.0000758.

22. Russell, T.W.; Hellewell, J.; Jarvis, C.I.; van Zandvoort, K.; Abbott, S.; Ratnayake, R.; working group, C.C..; Flasche, S.; Eggo, R.M.; Edmunds, W.J.; Kucharski, A.J. Estimatins the infection and case fatality ratio for coronavirus disease (COVID-19) using age-adjusted data from the outbreak on the Diamond Princess cruise ship, February 2020. Eurosurveillance 2020, 25, 2000256. doi:10.2807/1560-7917.ES.2020.25.12.2000256.

23. Verity, R.; Okell, L.C.; Dorigatti, I. Estimates of the severity of coronavirus disease 2019: A model-based analysis. The Lancet Infectious Diseases 2020, 20, 669. doi:10.1016/S1473-3099(20)30243-7.

24. Kermack, W.O.; McKendrick, A.G. A contribution to the mathematical theory of epidemics. Proceedings of the Royal Society A 1927, 115, 700. doi:10.1098/rspa.1927.0118.

25. Li, R.; Pei, S.; Chen, B.; Song, Y.; Zhang, T.; Yang, W.; Shaman, J. Substantial undocumented infection facilitates the rapid dissemination of novel coronavirus (SARS-CoV-2). Science 2020, 368, 489. doi:10.1126/science.abb3221.

26. Google. Google COVID-19 Comunity Mobility Reports, 2021.

27. Teralytics. Teralytics Matrix (Zürich, Switzerland), 2021.

28. Unacast. Unacast Social Distancing Scorecard, 2021.

29. Apple. Apple Mobility Trends, 2021.

30. Myers, J.L.; Well, A.D., Research Design and Statistical Analysis; Lawrence Erlbaum: London, 2003.

31. Rocha-Filho, T.M.; Mendesc, J.F.F.; Chow, C.C.; Phillips, J.C.; Cordeiro, A.J.A.; Scorza, F.A.; Almeidai, A.C.G.; AMoret, M. A data-driven model for COVID-19 pandemic – Evolution of the attack rate and prognosis for Brazil. Chaos, Solitons & Fractals 2021. In press, doi: 10.1016/j.chaos.2021.111359.

32. Lachassinne, E.; de Pontual, L.; Caseris, M.; Lorrot, M.; Guilluy, C.; Naud, A.; Dommergues, M.A.; Pinquier, D. SARS-CoV-2 transmission among children and staff in daycare centres during a nationwide lockdown in France: a cross-sectional, multicentre, seroprevalence study. The Lancet Child & Adolescent Health 2021, 5, 256. doi:10.1016/S2352-4642(21)00024-9.

33. Jrahmandad, J.; Lim, T.Y.; Sterman, J. Behavioral dynamics of COVID-19: estimating under-reporting, multiple waves, and adherence fatigue across 92 nations. SSRN 2021. Forthcoming, doi:10.2139/ssrn.3635047.

