## Supplemental Material for "A transnational and transregional study of the impact and effectiveness of social distancing for COVID-19 mitigation" (from another version of this preprint: version 1, 2021-09-05)

#### Materials and Methods

##### Effective reproduction number estimation

The effective reproduction number $R_t$ at the $i$-th day can be computed from the distribution for the generation time $w_i$ with $i$ the number of days between infections, as (*1*):

$$R_t(i) = \frac{I_i}{\sum_j w_j I_{i-j}}, \tag{S1}$$

with $I_i$ the number (or proportion) of infected individual at day $i$. The effective reproduction number can be estimated from the series of deaths by estimating the number of infected individuals from the number of deaths as:

$$I_i = \frac{1}{\overline{\theta}} \sum_j M_{i+j} u_j, \tag{S2}$$

where $u_i$ is the distribution of the number of days between first symptom and death (*3*), $M(i)$ the number of deaths in day $i$ and $\overline{\theta}$ is the average Infection Fatality Ration (IFR) (*2*), computed from the demographic structure in each locality. Having determined $I_i$ in this way we use Eq. (S1) to obtain $R_t$.

#### Spearman's rank-order correlation

To assess if the infection rate $\beta(t)$ is a (approximately) monotonous function of the mobility factor $M$, we use the Spearman's rank-order correlation $r_s(A, B)$ between two time series $A = (A_1, \ldots, A_{N_d})$ and $B = (B_1, \ldots, B_{N_d})$ of length $N_d$, where $A_i$ the value of the series at the $i$-th time value, and given by:

$$r_s[A, B] = 1 - 6\frac{\sum_{i=1}^{N_d} d_i^2}{N_d(N_d^2 - 1)}, \tag{S3}$$

such that $-1 \geq r_s \geq 1$ and $d_i$ is the difference in paired ranks of the two series $A$ and $B$, i. e. the difference in position of the $i$-th data point when ordered in ascending order in the data set. The coefficient $r_s$ measures the strength of how two variables are monotonically related, by an increasing or decreasing relation if $r_s > 0$ or $r_s < 0$, respectively.

In order to show the need for taking into account the decreasing number of susceptible individuals with time, we show in Fig. S1c and S1d the Spearman's correlation $r_s$ between the social isolation metric $M$ with the infection rate $\beta$, and between $M$ and the effective reproduction number $R_t$, for a period of $N_d = 150$ days, for the Los Angeles county in the United States. The dates in the Figure indicate the final day of the period considered. We observe that using $R_t$ as marker for the stage of the pandemic can lead to misleading results at later stages, as the proportion of susceptible individuals decays. Indeed, from Eq. (2) we see that the same value of $R_t$ can be associated to different values of the infection rate $\beta$, with the latter directly related to the circulation of the virus and its value reflecting more closely the different mitigation policies implemented.

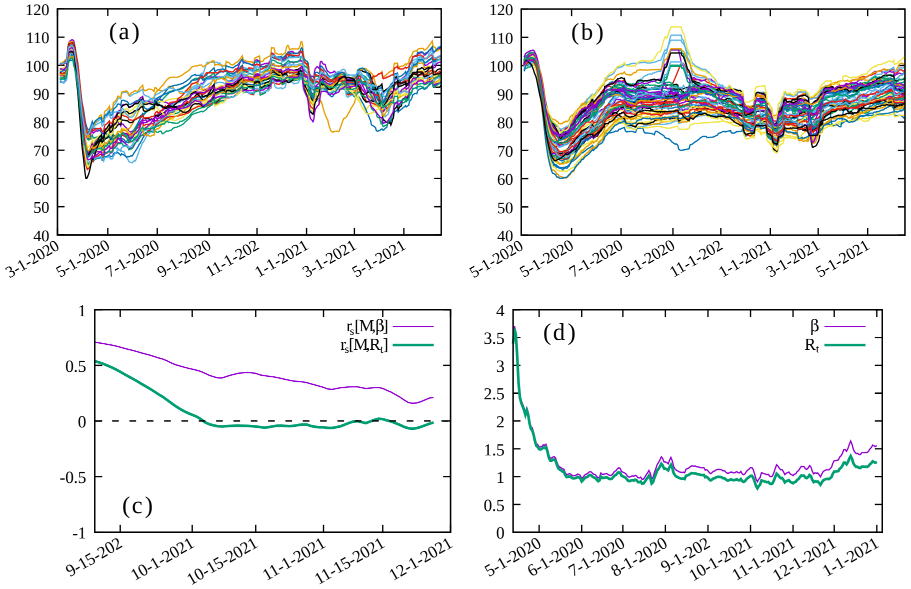


**Fig. S1.** Social distancing metric (social distancing index) $M$ in Eq. (1) for a) Brazil states and b) US states. c) Spearman's rank-order correlation $r_s[M, \beta]$ between the isolation index $M$ and the infection rate $\beta$ for the Los Angeles county in the United States, and $r_s[M, R_t]$ between $M$ and the effective reproduction number $R_t$. d) Time variation of $\beta(t)$ and $R_t$.

### References

1. C. Fraser. *PLoS ONE* **2**, e758 (2007).

2. R. Verity et al. *Lancet Infec. Dis.* **20**, 669-677 (2020)

3. T. W. Russell et al. *Euro Surveill.* **25**, 2000256 (2020).
